# The incremental value of computed tomography of COVID-19 pneumonia in predicting ICU admission

**DOI:** 10.1101/2021.01.08.20249041

**Authors:** Maurizio Bartolucci, Matteo Benelli, Margherita Betti, Sara Bicchi, Luca Fedeli, Federico Giannelli, Donatella Aquilini, Alessio Baldini, Guglielmo Consales, Massimo Edoardo Di Natale, Pamela Lotti, Letizia Vannucchi, Michele Trezzi, Lorenzo Nicola Mazzoni, Sandro Santini, Roberto Carpi, Daniela Matarrese, Luca Bernardi, Mario Mascalchi, COVID Working Group USL Toscana Centro and Azienda Ospedaliero-Universitaria Careggi, members of the Working Group USL Toscana Centro and Azienda Ospedaliero-Universitaria Careggi, Germana Allescia, Simone Busoni, Alessandra Bindi, Edoardo Cavigli, Diletta Cozzi, Giovanni Luca Dedola, Silvia Mazzocchi, Vittorio Miele, Chiara Pozzessere, Adriana Taddeucci, Adriano Viviani, Chiara Zini

## Abstract

**Rationale:** Triage is crucial for patient’s management and estimation of the required Intensive Care Unit (ICU) beds is fundamental for Health Systems during the COVID-19 pandemic.

**Objective:** To assess whether chest Computed Tomography (CT) of COVID-19 pneumonia has an incremental role in predicting patient’s admission to ICU.

**Methods:** We performed volumetric and texture analysis of the areas of the affected lung in CT of 115 outpatients with COVID-19 infection presenting to the Emergency Room with dyspnea and unresponsive hypoxyemia. Admission blood laboratory including lymphocyte count, serum lactate dehydrogenase, D-dimer and C-Reactive Protein and the ratio between the arterial partial pressure of oxygen and inspired oxygen were collected. By calculating the areas under the receiver-operating characteristic curves (AUC), we compared the performance of blood laboratory-arterial gas analyses features alone and combined with the CT features in two hybrid models (Hybrid radiological and Hybrid radiomics)for predicting ICU admission. Following a machine learning approach, 63 patients were allocated to the training and 52 to the validation set.

**Measurements and Main Results:** Twenty-nine (25%) of patients were admitted to ICU. The Hybrid radiological model comprising the lung %consolidation performed significantly (p=0.04) better in predicting ICU admission in the validation (AUC=0.82; 95%Confidence Interval 0.68-0.95) set than the blood laboratory-arterial gas analyses features alone (AUC=0.71; 95%Confidence Interval 0.56-0.86). A risk calculator for ICU admission was derived and is available at:https://github.com/cgplab/covidapp

**Conclusions:** The volume of the consolidated lung in CT of patients with COVID-19 pneumonia has a mild but significant incremental value in predicting ICU admission.

## Introduction

In January 2020 the World Health Organization declared Coronavirus disease (COVID-19) due to severe acute respiratory syndrome coronavirus 2 (SARS-CoV-2) a public health emergency of international concern. Until December 2020 it has caused more than 80 million of cases and more than 1.7 million of deaths worldwide (1).

About 15% of patients with COVID-19 pneumonia show a severe disease course requiring hospitalization and 5% eventual admission to an Intensive Care Unit (ICU) (2,3).

Prediction of ICU admission is crucial for patient’s management and forecasting the required number of ICU beds is fundamental for the resources allocation and organization of the health systems during the COVID-19 pandemic (4). Hence prediction of ICU admission has been frequently investigated in studies addressing diagnostic and prognostic models for COVID-19 (5). Among the variables potentially useful for such a purpose, clinical features yielded mixed results (6-12), while blood laboratory and arterial gas analysis features generally improved the overall prediction capacity (8,10,11,13-16).

Due to the non-specificity of findings, chest radiographs and computed tomography (CT) have no diagnostic role in patients with SARS-CoV-2 (17,18). Moreover the American College of Radiology and Society of Thoracic Radiology in the United States cautioned against their widespread use for assessment and monitoring disease course (18). However several studies in Asia, Europe and United States have indicated that chest radiographs and CT may have a role in the prediction of clinical evolution including need of ICU admission (19-25).

We hypothesized that CT-based quantitative analysis of the volume of the affected lung and its characterization in terms of texture analysis (26) might have an incremental role with respect to blood laboratory and arterial gas analysis results in predicting the patient’s admission to ICU. To explore this hypothesis following a machine learning approach we compared the predictive value of blood laboratory and arterial gas analysis features alone with those of two hyrbid models combining the same features with those derived from CT.

## Methods

The study received ethical approval (Protocol Number: 17260_oss) on May 19, 2020 by The Ethical Committee for Clinical Studies of the Central Tuscany Region. The study was performed between March 7^th^ and November 8^th^ 2020 at the Prato and Pistoia community hospitals in the Tuscany region of Italy, where overall 3-12% of hospital beds were allocated to ICUs in the study time period.

The study involved 208 outpatients with COVID-19 infection confirmed by positive nucleic acid test of real time-PCR in nasal-pharingeal swabs who presented to the Emergency Room (ER) and underwent immediate unenhanced chest CT because of dyspnea and non-responsive hypoxiemia. Laboratory data on admission included routine blood tests, serum lactate dehydrogenase (LDH), D-dimer and C-Reactive Protein (CRP) and lymphocytes count. Moreover in each patient the Horowitz (P/F) Index was calculated as the ratio between the arterial partial pressure of oxygen [PaO2] measured (in mmHg) by blood gas analysis and fraction of inspired oxygen [FiO2]. Patient’s age and gender and co-morbidities including history of arterial hypertension, diabetes, heart diseases and malignancies were annotated. Subsequent admission to the ICU and patient’s death were recorded as of November 13^th^, 2020.

Patients with CT images of low quality or incomplete blood laboratory or arterial blood gas analyses were excluded. Accordingly, data were analyzed in 115 of the initial 208 patients. Following a machine-learning approach and to avoid a “peeking” effect (27,28), 63 patients observed between March 7^th^ and April 21^st^, 2020 during the first wave of COVID-19 pandemic in Italy constituted the training set and 52 patients observed between August 18^th^ and November 8^th^ 2020 during the second wave constituted the validation set. Fig 1. shows the study flow-chart.The chest CT examinations were performed in Prato (n=88) on a Siemens SOMATOM Sensation (Germany) 64-rows of detectors scanner or in Pistoia (n=27)on an Optima CT660 GE Medical System (USA) 16-rows of detectors scanner. The patients were examined in supine position during inspiratory breath-hold or spontaneous breathing. CT acquisition parameters are detailed in an online data supplement. The CT images were transferred to a workstation implemented with the MIM Maestro software (MIM Software Inc.). Three Regions of Interest (RoIs) were automatically created from both lungs: Well-Areated Lung (WAL), which comprises the entire healthy tissue, Ground Glass Opacities (GGO), which includes areas showing ground glass density, and Consolidation (Consolid), which corresponds to areas of consolidated tissue. For segmentation of WAL and GGO, we used the *Region Growing tool* and threshold intervals were set from −950 Hounsfield Units(HU) to −700HU for WAL (21) and from −700HU to −250HU for GGO (29). For ConsolidRoI, a single expert radiologist with 20 years of experience in lung CT (M.B.) blind to the patient’s clinical, blood laboratory and arterial gas analyses results performed a manual editing of the segmentation results. An example is shown in Supplementary figure 1. The fraction of each RoI with respect to total lung volume was calculated. For image texture analysis, 107 radiomic features listed in Supplementary material table 1 were extracted using the 3DSlicer software (30) and the module radiomic (31). Image texture features were processed and analyzed by RadAR (Radiomics Analysis with R) (32). Features with duplicated ids and shape features (n=21) were excluded from downstream analysis.

**Table 1.** Clinical, blood laboratory, arterial gas analyses and CT results in patients with COVID-19 pneumonia. Continuous values are expressed as mean± standard deviation.

|  | Training (March 7- April 21, 2020 ) set |  |  | Validation (August 18 – November 8, 2020 ) set |  |  |
| --- | --- | --- | --- | --- | --- | --- |
|  | No ICU admission<br>(n = 44) | ICU admission<br>(n = 19) | P-value | No ICU admission<br>(n = 42) | ICU admission<br>(n = 10) | P-value |
| **Age, years** | 68 ± 12 | 66 ± 10 | 0.27 | 61± 3 | 72± 7 | **0.006** |
| **Gender** |  |  | 0.55 |  |  | 1 |
| *Male* | n = 32 (73%) | n= 12 (63%) |  | n = 23 (58%) | n = 7 (58%) |  |
| *Female* | n=12 (27%) | n = 7 (37%) |  | n = 17 (42%) | n = 5 (42%) |  |
| **Comorbodities** |  |  | 0.58 |  |  | 0.59 |
| 0 | n = 4 (11%) | n = 4 (27%) |  | n = 11 (30%) | n = 2 (20%) |  |
| 1 | n = 19 (51%) | n = 7 (47%) |  | n = 15 (41%) | n = 6 (60%) |  |
| 2 | n= 10 (27%) | n = 3 (20%) |  | n = 6 (16%) | n = 2 (20%) |  |
| >2 | n = 4 (11%) | n = 1 (7%) |  | n = 5 (14%) | n = 0 (0%) |  |
| **LDH (UI/L)** | 288 ± 127 | 371 ± 100 | **0.002** | 363 ± 135 | 400 ± 83 | 0.18 |
| **D-dimer (µg/mL FEU)** | 1.6 ± 2.6 | 2.2 ± 2.1 | 0.25 | 1.1± 1.0 | 2.0± 2.2 | 0.10 |
| **CRP (mg/dL)** | 9 ± 8 | 11± 9 | 0.35 | 6 ± 5 | 16 ± 10 | **0.0002** |
| **Lymphocytes(10<sup>3</sup>/uL)** | 1.1± 0.6 | 1.2 ± 1.2 | 0.63 | 1.2 ± 1.9 | 0.8 ± 0.4 | 0.21 |
| **P/F (mmHg)** | 236 ± 97 | 153± 66 | **0.001** | 243 ± 92 | 126 ± 47 | **0.00009** |
| **% Consolidation** | 5 ± 4 | 13 ± 10 | **0.0007** | 4 ± 4 | 8 ± 6 | **0.017** |
| **% Ground Glass** | 19 ± 16 | 29 ± 16 | **0.008** | 20 ± 15 | 26 ± 10 | 0.06 |
| **% Normal Lung** | 76± 18 | 58 ± 20 | **0.0009** | 76 ± 17 | 66 ± 11 | **0.026** |

Abbreviations: CRP= serum C-Reactive Protein; ICU= Intensive Care Unit; LDH= serum Lactate Dehydrogenase =; P/F= ratio between the arterial partial pressure of oxygen [PaO2] measured (in mmHg) by blood gas analysis and fraction of inspired oxygen [FiO2]

**Figure 1.**
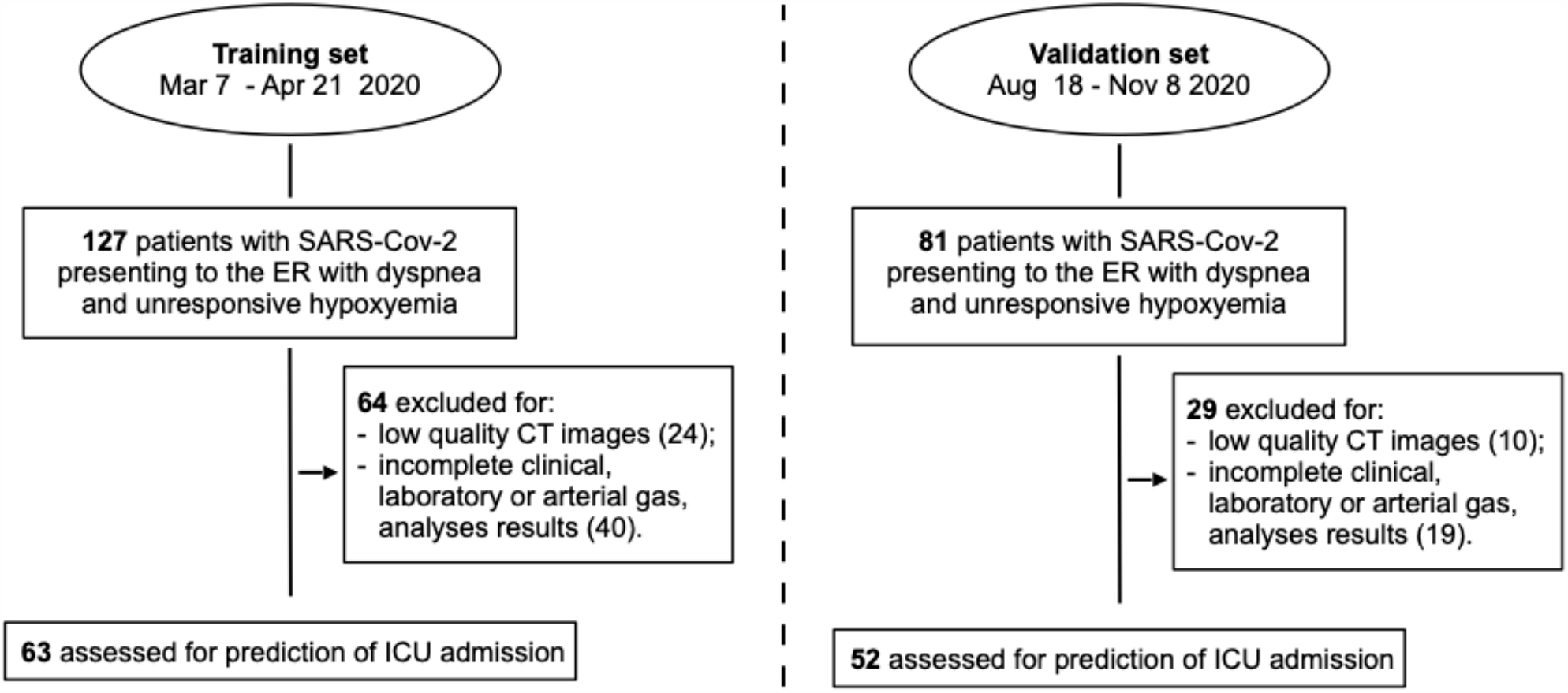
Study flow-chart. Abbreviations: CT = Computed Tomography; ER = Emergency Room; ICU = Intensive Care Unit; SARS-Cov-2 = Severe Acute Respiratory Syndrome Coronavirus 2

### Statistical Analysis and Model Construction

We used two-sided Wilcoxon-Mann-Whitney test to assess statistical significance of the differences between the ICU and non-ICU patients groups for parametric variables and Fisher’s Exact test for age and gender. Since co-morbidities can determine distortions in the admission to the ICU, especially when there is a relative shortage of dedicated beds, they were not included in the models. Models to predict ICU admission were built using binomial regression with GLMNET (33) on continuous variables, considering ICU admission as response variable (0=no, 1=yes). We selected GLMNET because, differently from other methods as multivariate random forest, it enables simultaneous selection of relevant features and parameter estimation. We built 5 models using age, blood laboratory features, the P/F ratio, the radiological and the radiomics features. The description of each feature is reported in Supplementary Table 1. To select relevant features for each model (feature selection), predictors showing nonzero coefficient at lambda.min - corresponding to the value of the regularization parameter lambda that gives minimum mean cross-validated error - were considered. To facilitate the applicability of our approach, GLMNET models were then rebuilt considering only model-specific selected features.

The performance of each model was assessed by calculating the area under the receiver operating characteristic curves (AUC) in the training and validation sets, using model probability as threshold parameter. All the analyses were performed using the R statistical programming language (https://cran.r-project.org/).

The confidence intervals of AUCs and the statistical significance of pairwise difference between AUCs were estimated by De Long’s tests implemented in the pROC R package (34).

### Web application

An interactive web application implementing the three best performing models (see below) was built using Shiny (https://CRAN.R-project.org/package=shiny).

## Results

Twenty-nine (19 of the training and10 of validation set) of the 115 included patients were admitted to ICU. The average interval between ER presentation and ICU admission was 1.9 days (range 1-22) in the training and 2.6 (range 1-5) in the validation set.

Table 1 details the distribution of age, gender, number of co-morbidities, blood laboratory and arterial gas analyses results and of those of the CT in the patients of the training and validation sets. In both sets the P/F index and %WAL were significantly lower and the %Consolid significantly higher in patients admitted to ICU. Comparing the data in the training and validation sets, only LDH was significantly higher (p=0.004) and the age lower (p=0.01) in the patients of the validation set who were not admitted to ICU.

Table 2 summarizes the considered features and those selected by the GLMNET. Age, LDH and the P/F ratio were selected as the best features both in the blood laboratory and arterial gas analyses model and in the Hybrid radiological model which also comprised %consolid. The Hybrid radiomics model comprised the P/F ratio, LDH, %consolid and 3 of the 86 considered texture features (see table 1 and supplementary table 1).

**Table 2.** Features *a priori* considered and features selected by GLMNET for their relevance in predicting ICU admission in five models.

| **Model** | **Features** | **Selected features (ICU admission)** |
| --- | --- | --- |
| Blood laboratory and arterial gas analyses | Age, LDH, D-dimer, PCR, Lymphocytes, P/F | Age, LDH, P/F |
| Radiological | %consolid, %ground glass, %normal lung | %consolid, %normal lung |
| Radiomics | 86 radiomic features (see Supplementary Table 1) | Dependence Non Uniformity, Small Dependence High Gray Level Emphasis, Large Dependence Low Gray Level Emphasis, Correlation, Interquartile Range, Total Energy, Run Variance, Large Area Low Gray Level Emphasis, Small Area Low Gray Level Emphasis |
| Hybrid radiological | Age, LDH, D-dimer, PCR, Lymphocytes, P/F, %consolid, %ground glass, %normal lung | Age, P/F, LDH, %consolid |
| Hybrid radiomics | Age, LDH, D-dimer, PCR, Lymphocytes, P/F, %consolid, %ground glass, %normal lung, 86 radiomic features | P/F, LDH, %consolid, Large Dependence Low Gray Level Emphasis, Run Length Non Uniformity, Low Gray Level Zone Emphasis |

Abbreviations: CRP= serum C-Reactive Protein; ICU= Intensive Care Unit; LDH= serum Lactate Dehydrogenase =; P/F= ratio between the arterial partial pressure of oxygen [PaO<sub>2</sub>] measured (in mmHg) by blood gas analysis and fraction of inspired oxygen [FiO<sub>2</sub>]; %consolid = percentage of consolidated lung; %ground glass = percentage of lung exhibiting ground glass opacities density; %normal lung = percentage of lung with normal density.

Fig. 2 shows the AUC of the model based on blood laboratory and arterial gas analyses features alone and of the two Hybrid models. The Hybrid radiological model performed better in predicting admission to the ICU in both the training (AUC=0.87; 95%Confidence Interval 0.77-0.97) and validation (AUC=0.86; 95%Confidence Interval 0.73-0.97) set as compared to the blood laboratory-arterial blood gas analyses features alone (training AUC=0.82; 95%Confidence Interval 0.68-0.95) (validation AUC=0.71; 95%Confidence Interval 0.56-0.86). The difference was significant (p=0.04) in the validation set. Also the Hybrid radiomics model performed better than the blood laboratory-arterial blood gas analyses features alone in the two sets, but the differences were not significant.

**Figure 2.**
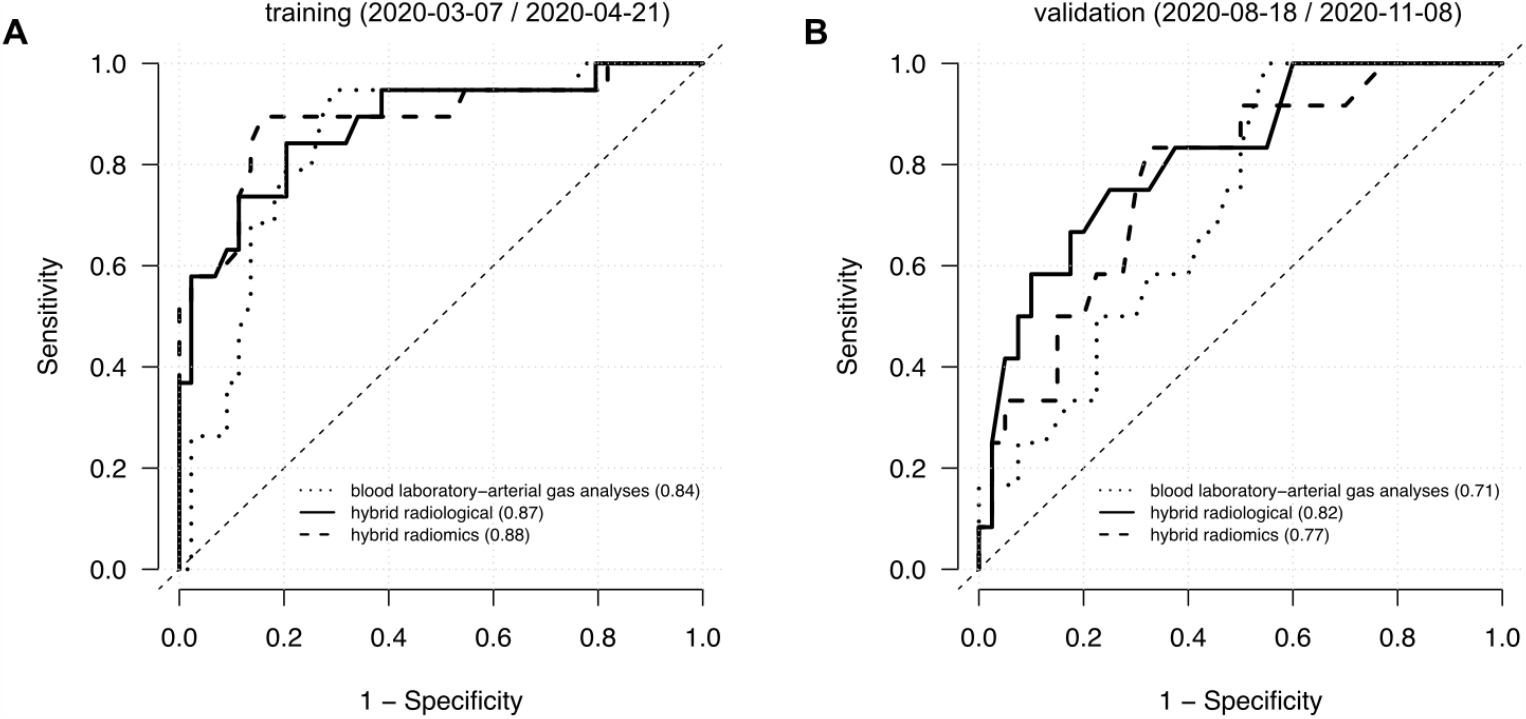
Performance of 3 models in predicting ICU admission. Receiving Operating Characteristic (ROC) curve analysis of the blood laboratory-arterial gas analyses features (dotted line), Hybrid radiological features (solid line) and Hybrid radiomics features (dashed line) in the training (A) and validation (B) sets. The values reported in parentheses refer to Area Under the ROC curves.

The models evaluating radiological and radiomics features alone selected two (%consolid and%WAL) and seven features(see supplementary table 1), respectively, but performed worse than the model evaluating blood laboratory and arterial gas analyses results in predicting ICU admission (see supplementary Fig. 2).

The distribution of the probability of ICU admission in the validation set (Supplementary Fig.3) indicates that no patient required ICU admission below the threshold of 0.25, 0.15 and 0.05 for the blood laboratory-arterial gas analyses model, Hybrid radiological and Hybrid radiomic models, respectively. In particular, in the Hybrid radiological model all patients with estimated probability below 0.20, corresponding to 16 (30%) of 52 patients, were all correctly classified and identified as a low-risk population without need of ICU admission.

The application to estimate the patient’s probability of ICU admission with the three best performing models is freely available at https://github.com/cgplab/covidapp under MIT license and allows users using the proposed models after insertion of the required model-specific features. Mortality rates were 14% (9/63) in the training and 21% (11/52) in the validation set.

## Discussion

Admission to ICU, where the most invasive and sophisticated treatments are carried out, is associated with a number of variables, including the evaluation of the patient’s clinical severity and evolution and the availability of ICU beds, but generally implies a severe structural and functional lung compromise, worse prognosis and increasing costs.

In this study we holistically combined blood laboratory, arterial gas analyses and CT results at ER presentation to predict ICU admission in patients with COVID-19 pneumonia. We demonstrated that the Hybrid radiological model combining CT estimation of volume of the consolidated lung with blood laboratory and arterial gas analyses features has a mild but significant incremental predictive value with respect to the model considering blood laboratory and arterial gas analyses features alone. This result is in line with and reinforces data from prior studies which evaluated the contribute of CT in predicting ICU admission in hybrid models (21,23-25).

For comparison with CT we considered several established blood biomarkers of severity of COVID-19 pneumonia including serum LDH, D-dimer, CRP and lymphocyte counts that can predict ICU admission (10,12,13,15,16,23). As well, we considered the P/F ratio which is a marker of non-responsive hypoxiemia in these patients (10,14,16).

Notably, in both our hybrid models the volume of consolidated lung, that is correlated with lung weight, was the best radiological biomarker instead than the volume of the well areated lung, that is the CT biomarker commonly used for Acute Respiratory Distress Syndrome (ARDS) (21,35,36).This is in line on the one hand with the observation that the pathological and radiological features of COVID-19 pneumonia are not typical of ARDS. In fact COVID-19 pneumonia along with diffuse alveolar damage and organizing pneumonia is characterized by a prominent vascular compromise justifying the observed disproportionate and non-responsive hypoxyemia (37,38) and the term “CARDS” (COVID-19 ARDS) (39) to label it. Moreover, according to Gattinoni et al. (40) consolidation and its extent characterizes two phenotypes of CARDS named type L (Low elastance, Low ventilation-to perfusion ratio, Low lung weight and Low recruitability) and type H(High elastance, High ventilation-to perfusion ratio, High lung weight and High recruitability) “which are best identified by CT”, involve different pathophysiological mechanisms and require different treatment options that, in case of type H, include intubation, positive end-expiratory pressure and extracorporeal membrane oxygenation that pertain to the ICU environment.

In our study, the Hybrid radiomics model including image texture features of the affected lung slightly (and not significantly) improved prediction of ICU admission as compared with blood laboratory and arterial gas analyses features, but performed worse than the Hybrid radiological model. Two prior studies reported a marginal incremental value of Radiomics for prediction of ICU admission as compared to volume estimation of the affected lung (24,25). Notably, since the Hybrid radiomics was the best performing model in our training set, but showed mild and non-significant advantage compared to blood laboratory and arterial gas features in the validation set, we speculate that Hybrid radiomics models might be more affected by overfitting as compared to the Hybrid radiological model. Overall, also considering that the pathological correlates of the CT texture features analysis are uncertain (26), we recommend estimation of the volume of lung consolidation and the Hybrid radiological model for triage of patients with COVID-19 pneumonia.

We recognize the following limitations of our study.

We performed a single centre study and assessment of the proposed models with data from other centres are required to verify their external validity. In our models we considered a large array of continuous variables in different domains, including D-dimer and P/F ratio which reflect the more characteristic physiopathological features of COVID-19 pneumonia(14,15,41) and are associated with worst prognosis (42), but discarded some potentially relevant categorical variables as gender, obesity and co-morbidities (43,44) which however are more closely linked with mortality than ICU admission. Moreover the recently described ABO blood-group system and genetic susceptibility loci (45) and some continuous variables as serum Interleukin-6 (23,46), ferritin and procalcitonin (16) were not available. Finally, we did not evaluate death as an outcome due to the small samples.However this would imply to consider treatments and other variables and was beyond the scope of the present investigation.

In conclusion, the combination of the volume of lung consolidation on CT at ER presentation has a mild but significant incremental value as compared to blood laboratory and arterial gas analyses results in predicting ICU admission. Inclusion of CT in the triage of patients with symptomatic COVID-19 pneumonia may have a practical value for individual patient’s management (possibly using the free application we developed) and help planning and organizing the Health Systems response to COVID-19 pandemic.

## Data Availability

Requests can be forwarded to
Maurizio Bartolucci or Matteo Benelli

https://github.com/cgplab/covidapp

## Figure Legends

**Supplementary Figure 1.**
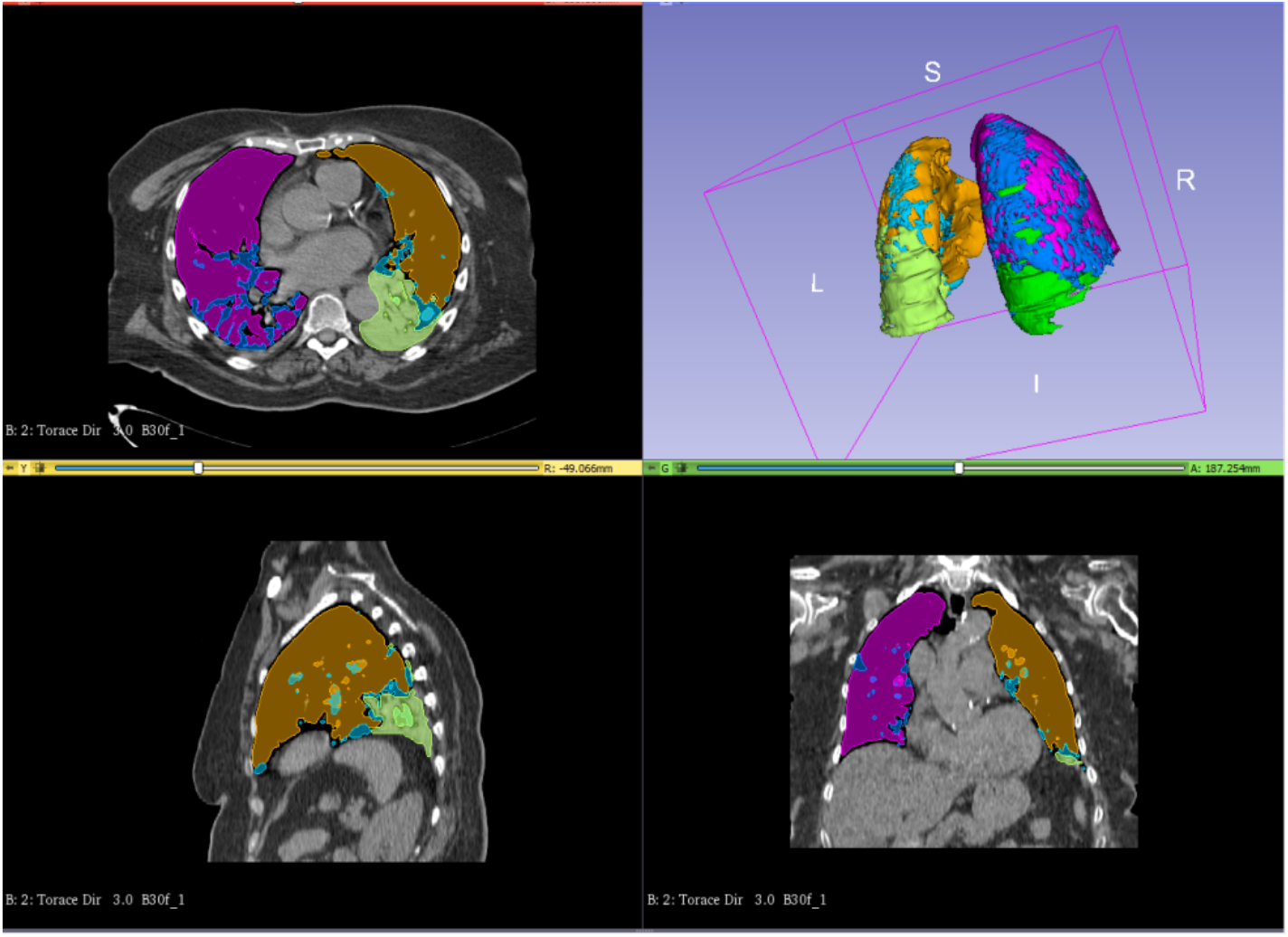
Example of segmentation of CT images. Well-Areated Lung (WAL) area are depicted in purple (right lung) and orange (left lung), Ground Glass Opacities (GGO) area are depicted in blue (right lung) and light blue (left lung) and Consolidation area (Consolid) are depicted in green (right lung) and light green (left lung).

**Supplementary Figure 2.**
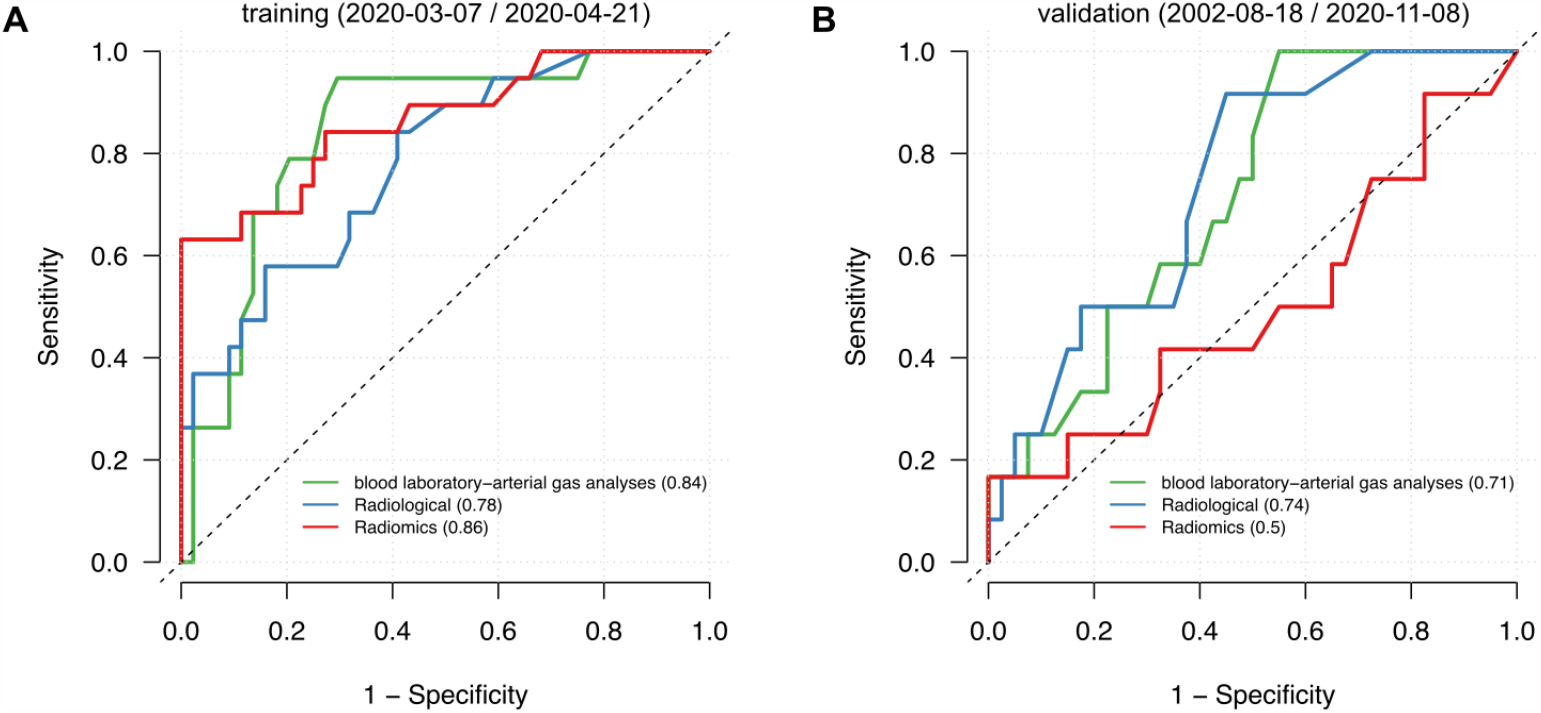
Performance of 3 simple models in predicting ICU admission. Receiving Operating Characteristic (ROC) curve analysis of the blood laboratory-arterial gas analyses features (green line), radiological features (sky blue line) and radiomics features (red line) in the training (A) and validation (B) sets. The values reported in parentheses refer to Area Under the ROC curves.

**Supplementary Figure 3.**
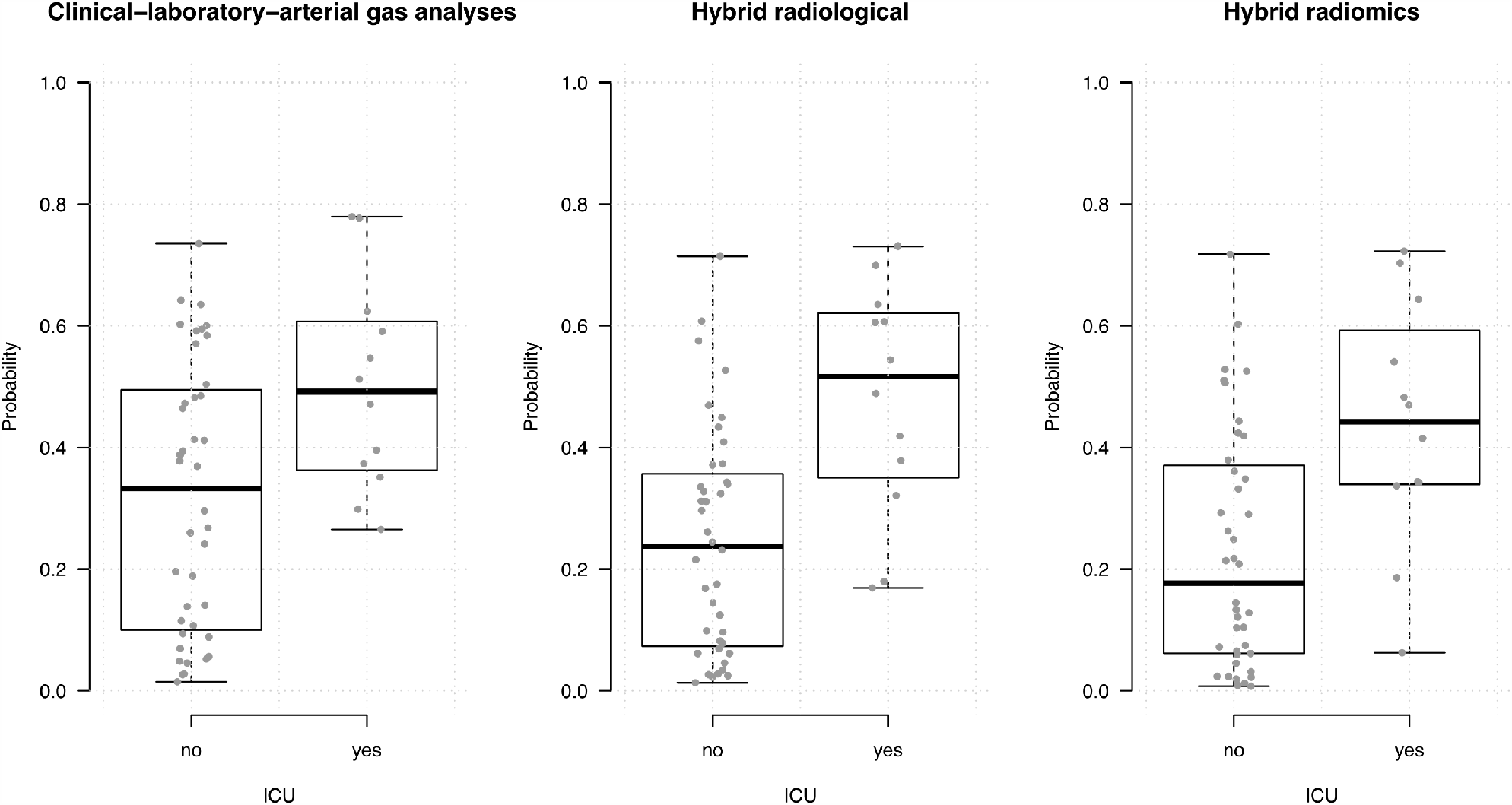
Box plots of the distribution of the probability of ICU admission estimated using the blood laboratory-arterial gas analyses (left), Hybrid radiological (middle) and Hybrid radiomics (right) models in the patients of the validation set who were not admitted (ICU=no) or required admission (ICU=yes) to ICU. No patient with estimated probability below 0.25 (laboratory-arterial gas analyses model), 0.15 (Hybrid radiology model) and 0.05 (Hybrid radiomic model) required ICU admission.

## On line repository material

### CT scanning parameters

They were set as follows: tube voltage 120 kV, tube current modulation from CareDose4D technology with quality reference mAs 150, pitch 1.4, slice thickness 0.6 mm in the Siemens scanner and tube voltage 120 kV, tube current modulation Smart mA technology, pitch 1.0, slice thickness 0.6 mm, in the GE scanner. Reconstruction filter used was B30 medium smooth for both the scanners and reconstruction was performed with slice thickness ranging from 2.5 and 3 mm in order to allow segmentation software to manage the amount of data in an appropriate time frame.

